# Prospective classification of functional dependence: Insights from machine learning and the Canadian Longitudinal Study on Aging

**DOI:** 10.1101/2024.07.15.24310429

**Authors:** Zachary M. van Allen, Matthieu P. Boisgontier

**Affiliations:** School of Rehabilitation Sciences, Faculty of Health Sciences, University of Ottawa, Canada; Perley Health Centre of Excellence in Frailty-Informed Care, Ottawa, Canada

**Keywords:** Aging, Exercise, Frailty, Memory, Pain, Muscle Strength, Physical Activity, Visual Acuity

## Abstract

**Purpose:** Functional dependence is a multifactorial health condition affecting well-being and life expectancy. To better understand the mechanisms underlying this condition, we aimed to identify the variables that best prospectively classify adults with and without limitations in basic and instrumental activities of daily living.

**Methods:** A filtering approach was used to select the best predictors of functional status from 4,248 candidate predictors collected in 39,927 participants aged 44 to 88 years old at baseline. Several machine learning models using the selected baseline variables (2010-2015) were compared for their ability to classify participants by functional status (dependent vs. independent) at follow up (2018-2021) on a training dataset (n=31,941) of participants from the Canadian Longitudinal Study on Aging. The best performing model was then examined on a test dataset (n=7,986) to confirm sensitivity, specificity, and accuracy.

**Results:** Eighteen candidate baseline variables were identified as the best predictors of functional status at follow up. Logistic regression was the best performing model for classifying participants by functional status and achieved balanced accuracy of 81.9% on the test dataset. The absence of functional limitations at baseline, stronger grip strength, being free of pain and of chronic conditions, being a female, having a drivers license, and good memory were associated with greater odds functionally independence at follow-up. In contrast, older age, psychological distress, walking slowly, being retired, having one or more chronic conditions, and never going for walks were associated with greater odds of functional dependence at follow-up.

**Conclusion:** Functional status can be best prospectively estimated by health condition, age, muscle strength, short-term memory, physical activity, psychological distress, and sex. These predictors can estimate functional status over 6 years ahead with high accuracy. This early identification of people at risk of functional dependence allows sufficient time for the implementation of interventions aiming to delay functional decline.

## Introduction

In 2020, the world’s population included 1 billion people aged 60 years or older. This number is expected to double by 2050. During the same period, the population aged 80 years or older is expected to triple (1). These demographic trends are associated with a greater number and proportion of people who experience difficulties with physical functioning (2,3), as manifested by a reduced ability to perform basic activities of daily living (ADL) (e.g., dressing, bathing, and walking) and instrumental activities of daily living (IADL) (e.g., shopping, managing one’s medications). These limitations have been shown to be a major contributor to increased early mortality (4) and decreased quality-adjusted life years (5). In addition, functional dependence is associated with patient financial barriers to care (6), caregiver burden (7,8), costly hospitalization (9), and long-term care home admission (10,11). As a result, preventing functional dependence is now considered a key component of healthy aging, which is defined as the process of developing and maintaining the functional ability that enables well-being in older age (12). However, while home-based interprofessional programs involving rehabilitation professionals have demonstrated efficacy in reducing ADL and IADL disability (13), healthcare systems and payers show only modest interest in the prevention of functional limitations, which may in part be due to the lack of direct reimbursement for preventive care related to physical function. Another possible explanation for this lack of interest is the difficulty in determining who should benefit from this preventive care.

The ability to predict which individuals are likely to become functionally dependent in the future is essential for identifying and intervening in high-risk cases. To enable and refine this prediction, studies have identified a range of variables associated with functional dependence. A 2007 systematic review of eight studies investigating early indicators of functional dependence found that age, lower functional status, disability, cognitive impairment, length of hospital stays, and depression were predictors of functional decline (14). Other research identified additional predictors such as physical activity (n = 12,860) (15), adverse childhood experiences (n = 25,775) (16), and disadvantaged childhood socioeconomic circumstances (n = 24,440) (17) in a 12-year European longitudinal study, as well as poor chewing ability in a 6-year Japanese study (n = 748) (18). In another Japanese sample (n = 1,523), walking speed, one-legged stance time as well as time to rise and walk were associated with a decline in functional independence (19). Additionally, lower socioeconomic status, unemployment, low fruit and vegetable consumption, and lower cognitive performance were associated with a greater risk of developing functional dependence over a four-year period in a Brazilian sample (n = 412) (20). Finally, age, chronic condition, and VO2max were associated with increased odds of becoming functionally dependent over the course of eight years in a Canadian sample (n = 297) (21).

However, some of these studies relied on small samples, most of them focused on a reduced number of candidate predictors, and none considered whether alternative modelling strategies would better fit the data. The machine learning approach is well suited to fill this gap, as it allows researchers to compare model performance between several algorithms (e.g., random forests, naive bayes) to select the model that is best able to predict or classify an outcome (22–25). To our knowledge, machine learning has not yet been applied to investigate the predictors that can prospectively classify functionally dependent and independent adults. However, machine learning has been applied to the prediction and classification of a condition closely related to functional dependence, namely frailty, an age-related state of vulnerability that affects multiple physiological systems (26). In a review that identified 217 frailty measurement tools, 52% included a functional limitation component (27), supporting that functional dependence and frailty have conceptual overlap. Studies using machine learning to classify frailty (28–31) have produced models with sensitivity (i.e., true positives) ranging from 74% to 99% and specificity (i.e., true negatives) ranging from 70% to 96%. In these models, factors such as income, age, the chair rise test, perceived health, chronic conditions, and balance problems demonstrated the strongest ability to classify frailty. However, it is unclear which indicators best classify functional limitations.

The objective of the present study was to use machine learning on a large cohort study to robustly identify the predictors that best prospectively classify functionally dependent and independent adults. Knowledge of such predictors would help identify individuals at higher risk of developing future functional limitations, inform the development of preventive interventions, and contribute to healthy aging.

## Methods

### Participants

Participants were recruited from the Canadian Longitudinal Study on Aging (CLSA), a nationally representative longitudinal study aimed at measuring the biological, societal, psychosocial, and physical factors related to healthy aging in Canada (32). Baseline data collection was conducted between 2010 and 2015 using two approaches: data collection from a ‘tracking’ cohort of participants was collected via hour-long computer-assisted phone interviews (n = 21,241) and data collection from a ‘comprehensive’ cohort via 90-minute in-person interviews in addition to a data collection site visits (n = 30,097). Additionally, a ‘maintaining contact questionnaire’ was administered by phone to both cohorts. The tracking cohort and comprehensive cohort were used as baseline data in our analyses. A follow-up assessment was from 2018 to 2021.

Participants in the CLSA were recruited through Canada’s provincial health registries, random-digit dialing, and the Canadian Community Health Survey on Healthy Aging (33,34). Exclusion criteria included: residents living in Canada’s three territories and First Nations reserves, full-time members of the Canadian Armed Forces, people living with cognitive impairments, and individuals living in institutions, including 24-hour nursing homes (32). Environmental measures from a separate study, the Canadian Urban Environmental Health Research Consortium (CANUE) (34), were linked with the CLSA dataset. The CANUE dataset contains multiple annual measures of greenness (e.g., tree canopy coverage), neighborhood characteristics (e.g., building density, public transportation), air quality (e.g., fine particulate matter, smoke exposure), and weather (e.g., local climate zones, land surface temperatures).

Data from 39,927 participants was partitioned into data used to train statistical models (training data, 80%; n = 31,941) and data used to test the predictive ability of the models (testing data, 20%; n = 7,986). Data used for classification was stratified by severity of ADL and IADL limitations so that equal proportions of older adults with low and high limitations at follow-up were present in both the training and the test datasets. All modeling on the training data used 10-fold cross-validation, which is generally considered best practice with machine learning (35). Resampling methods such as cross-validation enable the determination of how well a model works without using the test data.

### Dependent Variable

The primary dependent variable was limitations in ADL and IADL at follow-up, as assessed by a modified version of the Older Americans’ Resources and Services Multidimensional Functional Assessment Questionnaire (OARS) (36,37). Twenty questions were used to assess participants’ ability to complete seven ADL (i.e., can you dress and undress yourself without help, can you eat without help, can you take care of your own appearance without help, can you walk without help, can you get in and out of bed without any help or aids, can you take a bath or shower without help, do you ever have trouble getting to the bathroom in time). Twenty-one questions were used to assess seven IADL (i.e., can you use the telephone without help, can you get to places out of walking distance without help, can you go shopping for groceries or clothes without help, can you prepare your own meals without help, can you do your housework without help, can you take your own medicine without help, can you handle your own money without help).

The CLSA ‘Basic and Instrumental Activities of Daily Living Classification’ is based on the following scores: 1 (no limitation), 2 (mild limitation), 3 (moderate limitation), 4 (severe limitation), and 5 (total limitation) in ADL and IADL. The method used to derive this variable assigns extra weight to the ability to prepare meals and was scored to be similar to variables used in Statistics Canada’s Canadian Community Health Survey (33, 34). A binary variable from the Basic and Instrumental Activities of Daily Living Classification was computed which classified participants with no or mild limitation as functionally independent (coded 0), and participants with moderate, severe, or total limitation as functionally dependent (coded 1). As a sensitivity analysis, we classified participants with no limitation as functionally independent (coded 0) and participants with mild, moderate, severe, or total limitation as not fully functionally independent (coded 1) (Supplemental Material 2). This approach of dichotomising functional limitations has been used in previous research (38,39).

### Predictors Selection

Merging the variables from the tracking and comprehensive datasets resulted in an initial set of 4,248 variables, from which 3,574 variables were manually removed because they were single items of summary variables or outside the scope of the study (e.g., biomarkers). A total of 674 summary variables were considered as possible predictors of functional status (independent vs. dependent). Using a filtering approach (40), univariate logistic regressions were conducted between candidate predictors and functional status. A total of 630 variables were filtered out due to low associations with functional status. Subsequently, 26 variables were filtered out for containing redundant measures (e.g., multiple measures for age, income), using skip-logic questions, or through Least Absolute Shrinkage and Selection Operator (LASSO) reducing predictor estimates to 0 in logistic regression models, resulting in 18 predictor variables that were measured at baseline (Table 1): age, chronic conditions, driver’s license, grip strength, hip circumference, home care, limitations in ADL and IADL, memory (immediate and delayed recall), pain, perceived health, psychological distress, retirement, sex, urinary incontinence, visual acuity, walking frequency, and walking speed. Detailed descriptions of the variables used in analysis are presented in Supplemental Material 1.

**Table 1.** Descriptive statistics and logistic regression odds ratios.

| Baseline Predictors | Baseline Descriptive Statistics |  |  | Logistic Regression |
| --- | --- | --- | --- | --- |
|  | Full Sample<br>(n = 39,927) | Dependent at<br>Follow-up<br>(n = 1,147) | Independent at<br>Follow-up<br>(n = 38,780) | Odds Ratio for<br>Functional Dependence<br>at Follow-up |
| **Categorical Variables** | **Count (%)** | **Count (%)** | **Count (%)** | **Odds Ratios** |
| **Chronic Conditions** |  |  |  |  |
| No | 3,426 (8.8%) | 12 (1.0%) | 3,414 (8.8%) | 0.87 |
| 1 or more | 36,284 (90.9%) | 1,130 (98.5%) | 35,154 (90.6%) | 1.14 |
| Missing | 217 (0.5%) | 5 (0.4%) | 212 (0.5%) |  |
| **Driver's License** |  |  |  |  |
| Unrestricted | 37,422 (93.7%) | 816 (71.1%) | 36,606 (94.4%) | 0.91 |
| Other | 1,963 (4.9%) | 306 (26.7%) | 1,657 (4.3%) | 1.10 |
| Missing | 542 (1.4%) | 25 (2.2%) | 517 (1.3%) |  |
| **Home Care** |  |  |  |  |
| Receiving | 1,546 (3.9%) | 308 (26.9%) | 1,238 (3.2%) | 1.06 |
| Not Receiving | 38,346 (96.0%) | 836 (72.9%) | 37,510 (96.7%) | 0.94 |
| Missing | 35 (0.1%) | 3 (0.3%) | 32 (0.1%) |  |
| **Functional Limitations** |  |  |  |  |
| No | 36,467 (91.3%) | 578 (50.4%) | 35,889 (92.5%) | 0.79 |
| Mild | 2,980 (7.5%) | 373 (32.5%) | 2,607 (6.7%) | 1.00 |
| Moderate | 275 (0.7%) | 134 (11.7%) | 141 (0.4%) | 1.12 |
| Severe | 46 (0.1%) | 32 (2.79%) | 14 (0.04%) | 1.08 |
| Total | 19 (0.1%) | 14 (1.22%) | 5 (0.01%) | 1.04 |
| Missing | 140 (0.3%) | 16 (1.4%) | 124 (0.32%) |  |
| **Pain** |  |  |  |  |
| Pain free | 24,640 (61.7%) | 403 (35.1%) | 24,237 (62.5%) | .93 |
| Not pain free | 14,716 (36.9%) | 723 (63.0%) | 13,993 (36.1%) | 1.08 |
| Missing | 571 (1.4%) | 21 (1.8%) | 550 (1.4%) |  |
| **Perceived Health** |  |  |  |  |
| Poor | 572 (1.4%) | 101 (8.8%) | 471 (1.2%) | 1.11 |
| Fair | 2,893 (7.2%) | 265 (23.1%) | 2,628 (6.8%) | 1.17 |
| Good | 11,344 (28.4%) | 393 (34.3%) | 10,951 (28.2%) | 1.08 |
| Very Good | 16,726 (41.9%) | 293 (25.5%) | 16,433 (42.4%) | 1.00 |
| Excellent | 8,370 (21.0%) | 92 (8.0%) | 8,278 (21.3%) | 0.99 |
| Missing | 22 (0.1%) | 3 (0.3%) | 19 (0.1%) |  |
| **Retirement** |  |  |  |  |
| Completely retired | 16,783 (42.0%) | 866 (75.5%) | 15,917 (41.0%) | 1.16 |
| Partly retired | 4,545 (11.4%) | 87 (7.59%) | 4,458 (11.5%) | 1.00 |
| Not retired | 18,474 (46.3%) | 180 (15.7%) | 18,294 (47.2%) | 1.00 |
| Missing | 125 (0.3%) | 14 (1.2%) | 111 (0.3%) |  |
| **Sex** |  |  |  |  |
| Male | 19,307 (48.4%) | 458 (39.9%) | 18,849 (48.6%) | 1.10 |
| Female | 20,620 (51.6%) | 689 (60.1%) | 19,931 (51.4%) | 0.89 |
| **Urinary Incontinence** |  |  |  |  |
| Yes | 3,127 (7.8%) | 292 (25.5%) | 2,835 (7.3%) | 1.04 |
| No | 36,723 (92.0%) | 847 (73.8%) | 35,876 (92.5%) | 0.97 |
| Missing | 77 (0.2%) | 8 (0.7%) | 69 (0.2%) |  |
| **Walking Frequency** |  |  |  |  |
| Never | 6,013 (15.1%) | 353 (30.8%) | 5,660 (14.6%) | 1.12 |
| Seldom | 5,996 (15.0%) | 196 (17.1%) | 5,800 (15.0%) | 1.07 |
| Sometimes | 7,157 (17.9%) | 186 (16.2%) | 6,971 (18.0%) | 1.00 |
| Often | 20,186 (50.6%) | 384 (33.5%) | 19,802 (51.1%) | 0.95 |
| Missing | 575 (1.4%) | 28 (2.4%) | 547 (1.4%) |  |
| **Continuous Variables** | **Mean (SD)** | **Mean (SD)** | **Mean (SD)** | **Odds Ratio** |
| **Age (years)** | 62.0(9.9) | 70.9 (10.5) | 61.7 (9.7) | 1.83 |
| Missing | 0 | 0 | 0 |  |
| **Grip Strength (kg)** | 33.8 (11.4) | 26.2 (9.8) | 34.0 (11.4) | 0.81 |
| Missing | 16,334 | 566 | 15,768 |  |
| **Hip Circumference (cm)** | 104.1 (11.5) | 110.0 (15.5) | 104.0 (11.4) | 1.09 |
| Missing | 14,684 | 499 | 14,185 |  |
| **Memory (Immediate Recall)** | 6.0 (2.0) | 5.1 (2.0) | 6.1 (2.0) | 0.96 |
| Missing | 1,931 | 63 | 1,868 |  |
| **Memory (Delayed Recall)** | 4.3 (2.3) | 3.3 (2.1) | 4.4 (2.3) | 0.91 |
| Missing | 1,930 | 67 | 1,863 |  |
| **Psychological Distress** | 14.2 (4.4) | 17.0 (6.2) | 14.1 (4.4) | 1.22 |
| Missing | 15,118 | 488 | 14,630 |  |
| **Visual Acuity** | 0.3 (0.1) | 0.1 (0.2) | 0.03 (0.1) | 1.04 |
| Missing | 14,848 | 491 | 14,357 |  |
| **Walking Speed (sec)** | 4.2 (1.0) | 5.6 (1.8) | 4.2 (0.9) | 1.19 |
| Missing | 14,773 | 521 | 14,252 |  |

Note: Table presents the baseline data for all individuals in the sample, individuals who were functionally dependent at follow-up, and individuals who were functionally independent at follow-up. The odds ratios from the final logistic regression model are presented in the rightmost column. Missing data was not included in the model (imputation was used for missingness). Continuous variables were standardized in the models and odds ratios represent the odds for a 1 standard deviation increase in the predictor variable (i.e., the odds of being functionally dependent six years after baseline are 83% greater for a people who were 1SD greater than the mean age at baseline).

### Statistical Analyses

Modeling was conducted using the ‘tidymodels’ package (41), a meta-engine for machine learning models in R (42), which contains a collection of packages for modeling and machine learning in addition to workflow structures preventing common machine learning errors such as data leakage (43). A series of exploratory models were performed from which the best performing models were selected for subsequent interpretation (35). Only participants with complete data on the dependent variable (n = 39,927) were included in the analyses. Data imputation was performed for all predictor variables using bagged tree models, which can impute missing data for both numeric and categorical variables.

In machine learning, hyperparameters are set prior to the training process to guide the learning process. For example, hyperparameters include regularization strength in regression-based models, which prevents overfitting by adding a penalty to the models’ loss function that quantifies the error margin between a model’s prediction and the actual target value. Model/hyperparameter combinations refer to the various possible configurations of a model based on the values of its hyperparameters. Our analysis used a grid search approach to optimize hyperparameters and tested 125 model/hyperparameter combinations for classifying functional status.

During the training process, the values of the model parameters are determined. For the classification of functional status, the following packages and models were used: XGBoost (44), ranger (random forest) (45), klaR (naïve Bayes) (46), neural network (nnet) (47), and glmnet (logistic regression) (48). Numeric predictors for XGBoost, neural networks, and logistic regressions were normalized. Categorical predictors were encoded into numerical data using one-hot encoding (i.e., category levels are represented independently as separate binary variables without comparison to a reference group). Hyperparameters for each model were determined with a grid search methodology using the ‘tune’ package (43) and the best performing models were selected for further fitting and evaluation. The models selected for this study mirror those used in similar predictive modelling studies (28–31).

During the model training and testing processes, model performance was evaluated through the level of misclassification on out-of-sample data using a standard confusion matrix (22), which represents different combinations of predicted and actual values forming the basis of metrics for precision, specificity, and accuracy. In addition, area under the curve (AUC) of the receiver operating characteristic (ROC) curve was used to represent how well the model can distinguish between functional statuses (35). A 50% AUC indicates random guessing, and 100% indicates perfect performance. Here, this score was interpreted as was interpreted as: ≥ 60%, poor; ≥ 70%, fair; ≥ 80% good; ≥ 90%, excellent (49). Specificity refers to the proportion of true negative, sensitivity refers to the proportion of true positives, and accuracy is the proportion of true positive and negative results among the total cases examined. By default, binary classification models use a threshold of 0.50 to predict if a given case should be classified as 0 (functionally independent) or 1 (functionally dependent); the optimal threshold value which balances specificity and sensitivity was computed for each model.

## Results

### Descriptive Results

A total of n = 39,927 participants had reported their functional status and were retained in the final sample, which included English (n = 32,298) and French speakers (n = 7,629), 20,620 females (51.6%). Age ranged from 44 to 88 years at baseline. Detailed descriptive statistics of the sample’s characteristics are presented in Table 1.

When comparing the baseline characteristics between functionally independent and dependent participants at follow-up, several differences between groups were observed. For example, 92.5% of participants classified as independent at follow-up reported no impairments at baseline while 50.4% of those classified as dependent at follow-up reported no impairments at baseline. Additionally, dependent participants reported a greater rate of chronic conditions at baseline (98.5% vs. 90.6%), greater pain (63.0% vs 36.1%), not having a driver licence (26.7% vs. 4.3%), higher rates of urinary incontinence (25.5% vs. 7.31%), poorer perceived health (8.8% vs. 1.2%), higher retirement rates (75.5% vs. 41.0%), weaker grip strength (26.2 ± 9.8 kg vs. 34.0 ± 11.4 kg), and older age (70.9 ± 10.5 years vs. 61.7 ± 9.7 years). The mean number of years between baseline and follow-up was 6.3 years (range: 3.9 to 9.6 years).

### Model Performance

Models classifying participants with no or mild limitation as functionally independent participants and participants with moderate, severe, or total limitation as functionally dependent provided the strongest accuracy. Analyses using the alternate classification scheme are presented in Supplemental Material 2.

The best two models demonstrated approximately equivalent performance on the training data: logistic regression (AUC = 89.2%, accuracy = 97.3%, specificity = 99.7%, sensitivity = 13.0%) and XGBoost (AUC = 89.1%, accuracy = 97.4%, specificity = 99.8%, sensitivity = 12.8%) (Table 2, Figure 1). When the threshold of 0.50 was adjusted to optimise a balance between sensitivity and specificity, model performance increased to above 80% sensitivity and 80% specificity for each of these two models (Table 2). When considering the overall classification accuracy using balanced sensitivity and specificity, logistic regression was the best performing model with 82.0% accuracy (vs. 81.7% for the XGBoost model). Model performance for logistic regression on the test data was approximately equivalent to the training data (balanced accuracy = 81.9%, specificity = 79.0%, sensitivity = 84.7%), suggesting the model did not overfit on the training data.

**Figure 1.**
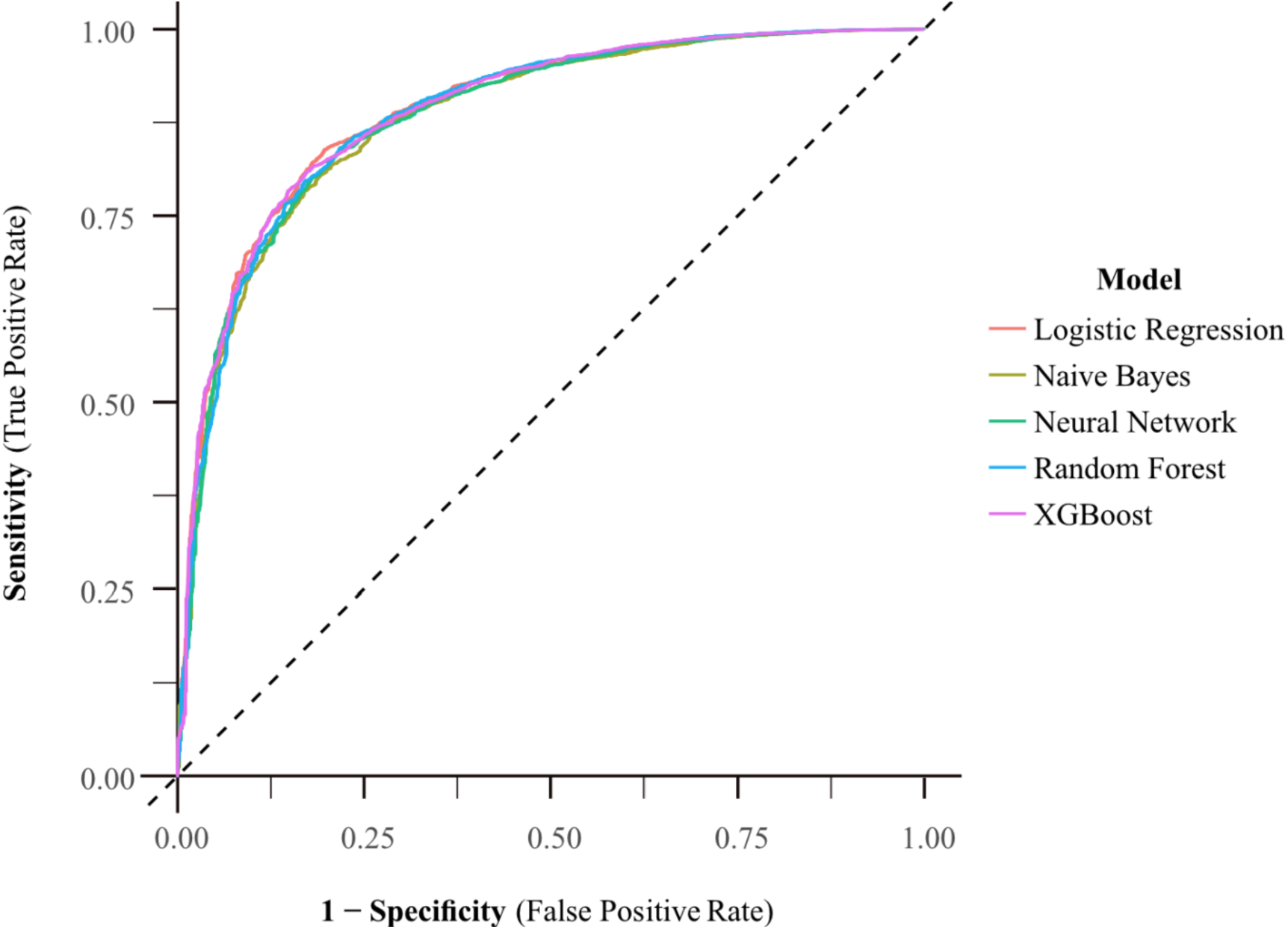
Receiver operating characteristic (ROC) curves of models classifying functional status at follow-up (dependent vs. independent) using data from the 18 baseline predictors of the training dataset (n = 31,941).

**Table 2.** Model performance on training and test data.

| Model | AUC | Accuracy | Sensitivity | Specificity | Balanced<br>Threshold | Balanced<br>Sensitivity | Balanced<br>Specificity | Balanced<br>Accuracy |
| --- | --- | --- | --- | --- | --- | --- | --- | --- |
| **Training Data (n = 31,941)** |  |  |  |  |  |  |  |  |
| Logistic<br>Regression | 89.2% | 97.3% | 13.0% | 99.7% | .9711 | 81.9% | 82.2% | 82.0% |
| XGBoost | 89.1% | 97.4% | 12.8% | 99.8% | .9753 | 82.5% | 81.0% | 81.7% |
| Random<br>Forest | 88.8% | 97.2% | 0.0% | 100.0% | .9660 | 82.8% | 79.7% | 81.3% |
| Naïve Bayes | 88.1% | 94.5% | 45.7% | 95.9% | .9924 | 81.2% | 80.1% | 80.6% |
| Neural<br>Network | 88.4% | 97.3% | 13.0% | 97.3% | .7212 | 82.1% | 79.9% | 81.0% |
| **Test Data (n = 7,986)** |  |  |  |  |  |  |  |  |
| Logistic<br>Regression |  |  |  |  | .9711 | 84.7% | 79.0% | 81.9% |

### Best Classifiers

The predictor that best classify functionally independence at follow-up was having no functional limitations at baseline with participants reporting no limitations at baseline being 21% more likely to be functionally independent at follow-up (OR = 0.79) (Figure 2). Additionally, grip strength (OR = 0.81), the absence of chronic conditions (OR = 0.87), being female (OR = 0.89), having an unrestricted driver’s license (OR = 0.91), higher scores on a delayed recall memory test (OR = 0.96) at baseline were all associated with greater odds of being functionally independent at follow-up. Note that continuous variables were standardized and ORs represent the odds for a 1SD increase in the predictor variable. Conversely, the following baseline variables were associated with higher odds of being dependent at follow-up: older age (OR = 1.83), psychological distress (OR = 1.22), longer times to complete a walking task (OR = 1.19), being completely retired (OR = 1.16), having one or more chronic conditions (OR = 1.14), and ‘never’ engaging in weekly walks (OR = 1.12).

**Figure 2.**
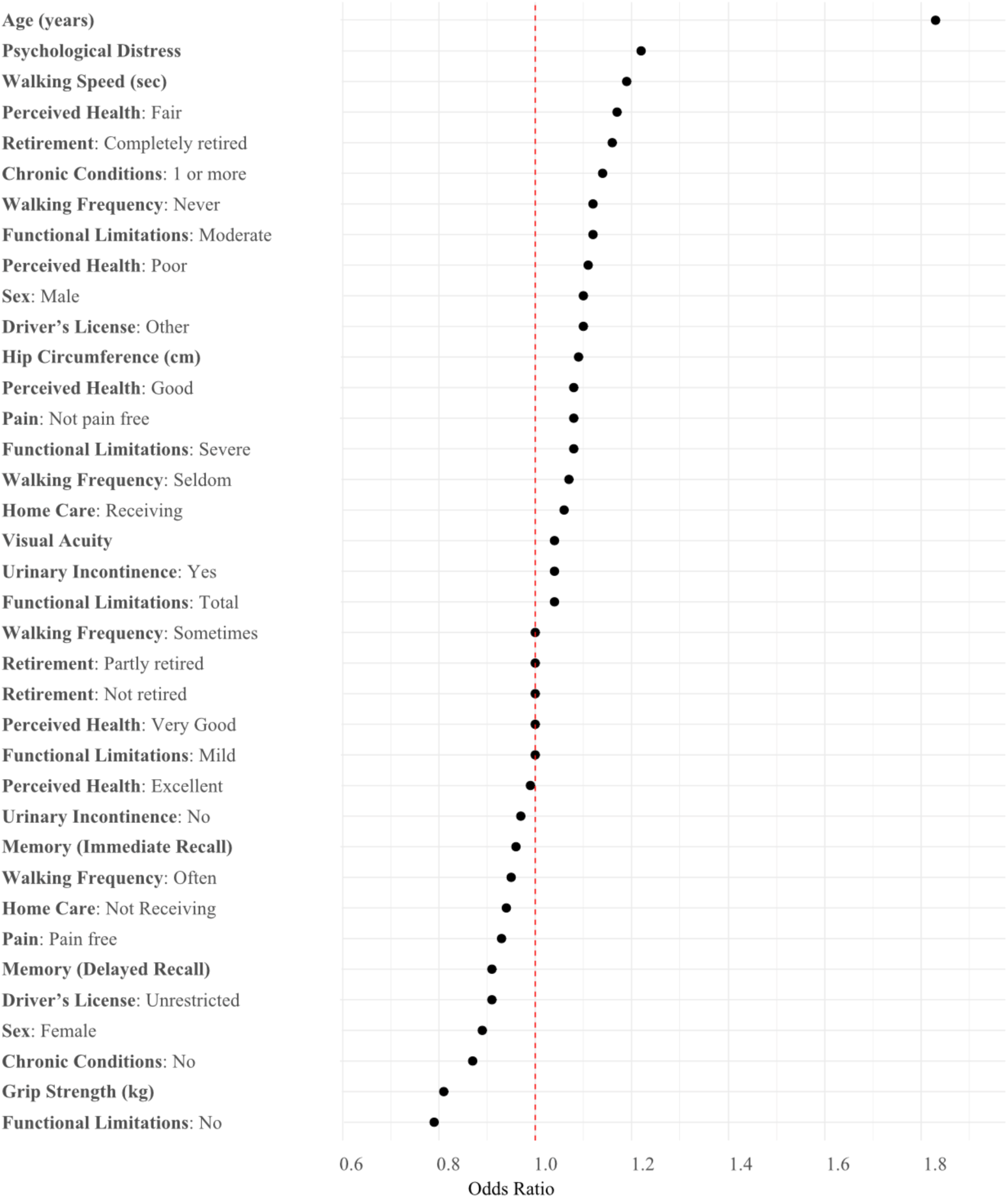
Forest plot visualising odds ratios of baseline variables most strongly associated with functional dependence at follow-up. Notes: Continuous variables (e.g., age) were standardized and ORs represent the odds for a 1SD increase in the predictor variable. Confidence intervals are not available for regularized logistic regression models. Regularization techniques (like Lasso or Ridge regression) introduce a bias into the parameter estimates by shrinking the coefficients toward zero. This is done to prevent overfitting and improve generalization, especially when dealing with high-dimensional data. Because the coefficients are intentionally biased, the usual methods of estimating confidence intervals (based on unbiased estimates) become less accurate or invalid.

## Discussion

This study applied machine learning techniques to data from a large cohort study (n = 39,927) to identify the variables from baseline assessments (2011-2015) that best classify functionally dependent and independent adults at follow-up (2018-2021).

Eighteen variables were identified as the best predictors of functional status: age, chronic conditions, driver’s license, grip strength, hip circumference, home care, limitations in ADL and IADL, memory (immediate and delayed recall), pain, perceived health, psychological distress, retirement, sex, urinary incontinence, visual acuity, walking frequency, and walking speed. Using these variables, the best model showed a high proportion of true negatives (79%) and true positives (85%), demonstrating high accuracy in correctly identifying functionally dependent and independent adults (82%). The following baseline variables were the best classifiers of dependent and independent adults at the follow-up assessment: perceived health, age, grip strength, chronic conditions, functional limitations, memory, walking frequency, and psychological distress. None of the environmental variables from the CANUE dataset (e.g., greenness, neighborhood characteristics, air quality, and weather) showed strong evidence of an association with functional status This was surprising given the hypothesized link between environmental quality and frailty (50). However, it is possible that longer exposures to sub-optimal environments may be required for effects to become identifiable.

Our model outperforms some recent models focusing on frailty (28,29), which is closely related to, but distinct from, functional dependence (27). For example, Aponte-Hao et al. (28) used two years of electronic medical records from Canadian primary care practices to predict frailty with the Rockwood Clinical Frailty Scale (51) and obtained 78% sensitivity and 74% specificity. Tarekgen et al. (29) used one-year of data from the Piedmontese Longitudinal Study to predict five indicators of frailty (mortality, urgent hospitalization, disability, fracture, and emergency admission) with sensitivity ranging from 75% to 81% and specificity from 70% to 80%. While comparison with these studies is difficult due to differences in the assessed concepts (frailty vs. functional status), variability in the time between assessments, differences in national contexts, and the nature of the available predictors in the dataset, our model obtained reasonable performance with greater distance between data measurements and fewer prediction variables (n = 18) than previous studies [n = 75 (28); n = 58 (29); n = 27 (30)]. This latter result is clinically important as this parsimony facilitates future work of personalized prediction of those at risk of becoming functionally dependent.

While our models outperformed some previous work, higher model performance has been reported in the frailty literature (30,31,52). For example, Leme and De Oliveira (31) predicted frailty using four years of data from the English Longitudinal Study of Aging to predict frailty using the Fried phenotype measure (53) and obtained 83% sensitivity and 88% specificity with the strongest predictors of frailty including age, the chair rise test, perceived health, and balance problems. Interestingly, while age and perceived health were among the strongest predictors in our analysis, the chair-rise test, income, and balance problems were not. These latter variables may be related to dimensions of frailty that are unrelated to functional dependence. National context (Canada vs. Brazil) may partly explain the discrepancy with household income. The inclusion of several physical fitness measures in the present study (e.g., walking frequency, walking speed, grip strength) may have accounted for the relationship between physical capacity and frailty. In another example, Koo et al. (30) used 27 predictors in the Korean Frailty and Aging Cohort, over a one-year period, to predict scores on the Fried Frailty Index and achieved a score in which sensitivity and specificity contribute equally (F1 score) of 95.3%. Notably, these higher performing models were obtained with shorter intervals between data collection waves [6.3 years on average in the present study vs. one year (29) and four years (30)]. When classification is performed in a single observational event (e.g., using a motion capture system during body movements to predict frailty), classification performance can increase to 97.5% (52). However, it is important to keep in mind that although greater classification performance can be achieved when the time between predictor and outcome measures is shorter, the purpose of modeling studies is not to assess health status. The value of modeling studies lies in their ability to identify as many years in advance as possible who is at risk of becoming frail or functionally dependent, allowing sufficient time to intervene and delay or even prevent this functional decline. Future modelling studies using functional limitations as the outcome variable are needed to make more accurate comparisons between model performance.

### Strengths and Limitations

This study has several strengths including (i) the use of a large cohort study, (ii) the investigation of functional limitations that fills a gap in the literature, (iii) the exploration of predictors of functional limitations at multiple levels (individual, social, environmental), and (iv) similar high accuracy across the tested models, suggesting well-selected variables and robust results. Several limitations should also be noted. First, the length of time between baseline and follow-up assessments varied due to the nature of CLSA data collection and it is unclear whether sufficient time between timepoints was sufficient for causal relationships to manifest. Second, a strong class imbalance was present in the data with only 2.9% of participants classified as functionally dependent. Synthetic minority oversampling techniques (SMOTE) (54) could have been used to generate a balanced dataset with synthetic data. However, this technique is not without its limitations and can introduce noise into a model (55). In our analyses, we opted to balance the proportion of functionally dependent adults in the training and test dataset without artificially creating data.

## Conclusions

To our knowledge, we present the first study that identifies and models the variables that best prospectively classify functionally dependent and independent adults. Results from the comparisons of predictive models suggest that regularized logistic regression outperforms more complex statistical models and is suitable for the task of classifying functional limitations. The identified predictors can predict functional status more than 6-year in advance (range 3.9 to 9.6 years) with high accuracy. Because functional ability is a key component of healthy aging (12), preventing its decline should be a priority to healthcare providers, especially in the context of an aging population (1). Early identification of individuals at risk allows healthcare professionals sufficient time to implement interventions aimed at delaying or preventing functional decline, and to refer patients to the appropriate specialists when needed, before functional decline develops.

While all healthcare providers can incorporate such preventive care into their treatment plans, rehabilitation professionals play a central role in maintaining and optimizing functional abilities. In addition, predictors such as grip strength, walking speed and frequency, and pain management fall within their scope of practice. Screening for these predictors should be considered when assessing patients, especially older adults, to allow for early intervention. Patient education regarding the identification of predictors of functional decline and the importance of addressing them should not be overlooked, as this may help motivate patients to actively take part in their care. In short, prospective identification and early intervention of functional dependence could have an important impact on preventing loss of autonomy, reducing significant costs to individuals and the healthcare system, as well as reducing caregiver burden and long-term care admissions.

## Declarations

### Data and Code Availability

In accordance with good research practice (56) the R scripts are publicly available in Zenodo (57). This research has been conducted using the CLSA dataset (Baseline Tracking Dataset version 4.0, Baseline Comprehensive Dataset version 7.0, Follow-Up 2 dataset version 1.1) under application number 2304007. This dataset is available for researchers who meet the criteria (www.clsa-elcv.ca).

### Authorship Contribution Statement

Based on the Contributor Roles Taxonomy (CRediT) (58) individual author contributions to this work are as follows:

- Zachary van Allen: Conceptualization; Methodology; Formal Analysis; Writing (Original Draft); Writing (Review and Editing).
- Matthieu Boisgontier: Conceptualization; Methodology; Writing (Original Draft); Writing (Review and Editing); Supervision (ZMvA); Project Administration; Funding Acquisition.

### Reporting Guidelines

This manuscript conforms to the STROBE guidelines for observational studies (59).

